# Impact of the COVID-19 Pandemic on Adult Asthma-Related Healthcare Utilization

**DOI:** 10.1101/2025.09.19.25336168

**Authors:** Lizbeth F. Gómez, Kimberly D. Lactaoen, Patrick K. Gleeson, Alana Schreibman, Jason D. Christie, Andrea J. Apter, Rebecca A. Hubbard, Gary E. Weissman, Blanca E. Himes

## Abstract

**BACKGROUND:** The COVID-19 pandemic prompted unprecedented changes to chronic disease self-management and healthcare systems worldwide, including shifts in access to services and medications. While children with asthma had decreased exacerbations and healthcare encounters during 2020, the impact of lockdowns on adults with asthma, who faced different challenges during the pandemic than children, are less understood.

**OBJECTIVE:** We sought to characterize changes in adult asthma-related healthcare utilization during the COVID-19 pandemic in 2020 versus prior (2017-2019) and subsequent (2021-2024) years by leveraging electronic health record (EHR) data from a large, multi-hospital health system in a major US city.

**METHODS:** We conducted a retrospective EHR database study of 42,242 adults with asthma who received care at Penn Medicine from 2017 to 2024. We analyzed weekly encounter counts across five encounter types (refill, telemedicine, telephone/audio, outpatient, emergency encounters) and prescriptions for short-acting beta agonists (SABA), inhaled corticosteroids (ICS), and oral corticosteroids (OCS). Generalized linear models assessed changes in asthma-related encounter rate in pandemic (2020) and post-pandemic (2021-2024) periods relative to pre-pandemic (2017-2019). We stratified on weekly intervals that captured transitional timepoints in healthcare utilization in 2020 (Weeks 1-8, 9-18, and 19-52).

**RESULTS:** In 2017-2019, there were on average 397 weekly visits for asthma at Penn Medicine; in 2020, the weekly average increased to 481. This change was driven primarily by a surge during the lockdown weeks in refill and telemedicine encounters by 123% and 36,445%, respectively and by a decrease in outpatient visits by 65%. During the lockdown weeks in 2020, asthma related prescriptions of SABA and ICS prescriptions increased 73% and 43%, respectively, compared to pre-pandemic years, while OCS prescriptions decreased by 5%. White patients showed earlier healthcare-seeking responses than other racial groups. Changes persisted in post-pandemic years as the average of weekly asthma-related visits was 445 in 2021-2024. Telemedicine remained 38-76 times higher than pre-pandemic baseline, refills doubled compared to 2017-2019 levels, and outpatient visits remained 35-43% below pre-pandemic levels.

**CONCLUSION:** COVID-19 transformed adult asthma care delivery and led to sustained increases in virtual care and medication refills potentially due to virtual care compensating for reductions in traditional outpatient encounters.

## INTRODUCTION

The COVID-19 pandemic brought about unprecedented changes to healthcare systems worldwide. Public health mandates, aimed at curbing viral transmission, both mitigated the spread of COVID-19 and prompted significant shifts in how patients engaged with healthcare services. Early in the pandemic, there was increased prescription and dispensing rates for medications, which were attributed to “stockpiling” behavior as individuals attempted to secure essential supplies amid uncertainties.^1–3^ Similarly, there was a transformation in healthcare delivery as telehealth became widespread.^4^ Telehealth services, defined as the remote provision of healthcare using information and communication technologies, rapidly evolved from a niche offering to a mainstay of clinical practice. One U.S. study found that the proportion of ambulatory visits using telehealth services increased steeply from 0.3% in 2019 to over 20% in 2020.^1^ Concurrent with changes in healthcare delivery were shifts in individual behaviors in response to mandated public health precautions, including the use of personal protective equipment and social distancing, both of which decreased exposures to particulates and subsequently, may have modified associated asthma incidence.^5–8^

Asthma is a chronic inflammatory disorder of the airways that requires monitoring, medication adjustments, and timely intervention to prevent and treat episodes of worsening symptoms that are commonly treated with oral corticosteroids. The COVID-19 fundamentally transformed healthcare delivery models with widespread changes in access, care continuity, and medication dispensing that disrupted routine healthcare utilization.^9^ For adults with asthma, these disruptions occurred alongside complex and potentially competing factors: reduced exposure to ambient pollutants, asthma triggers, and decreased viral transmission may have improved disease control, while barriers to seeking routine care may have led to delayed or foregone treatment. However, the magnitude, duration, and clinical implications of these changes in adult populations remain unclear.^10^ To address this knowledge gap, we measured patterns of adult asthma-related healthcare utilization using electronic health record (EHR) data from a large, multi-hospital health system spanning encounters from 2017 to 2024.

## METHODS

### Study design and population

We performed a retrospective database study of 42,242 unique patients who received healthcare for asthma at Penn Medicine, a large health system that serves the greater Philadelphia area. We included patients who were 18 years of age or older, had encounters that occurred between January 1, 2017, to December 31, 2024, and had at least one International Classification of Disease, Tenth Revision (ICD-10) code for asthma (J45^*^). Encounters were limited to refill, telephone, telemedicine, outpatient, and emergency types from clinical sites that contributed data throughout the 8-year study period. Patient data included sex (Female, Male), race (American Indian/Native Alaskan [AIAN], Asian, Black, Native Hawaiian/Pacific Islander [NHPI], White, and Other), ethnicity (Hispanic/Latino, non-Hispanic/Latino, Unknown/Other), age at first encounter (18-24, 25-34, 35-44, 45-54, 55-64, 65-74, 75-84, 85+), body mass index (BMI; Not Overweight or Obese, Overweight, Obesity Class I, Obesity Class II, Obesity Class III, Unknown/Other), insurance type (Medicaid, Medicare, Private, Unknown/Other), smoking status (Current Smoker, Former Smoker, Never Smoked, Unknown/Other), Elixhauser score^11^ (<0, 0, 1-9, 10+) and asthma-related medication prescriptions (inhaled corticosteroids [ICS], oral corticosteroids [OCS] and short-acting beta agonists [SABA]). Patients with cystic fibrosis as determined by ICD-10 code (E84^*^) and medications for cystic fibrosis (i.e., transmembrane conductance regulator [CFTR] modulators, dornase alfa, and mannitol), and those with missing race or reported “other” for sex were excluded from the analysis. This study was approved by the University of Pennsylvania Institutional Review Board under protocol number 824789. Formal consent was not obtained, as this research was conducted under a granted waiver of HIPAA Authorization.

### Asthma healthcare utilization

Asthma healthcare utilization was defined as the weekly number of asthma-related encounters for refill, telemedicine, telephone, outpatient and emergency encounter types as recorded in the EHR. Counts represented the total volume of asthma-related encounters within Penn Medicine and reflected both changes in individual patient care-seeking behavior and changes in the patient population served by the system over time. An encounter was classified as asthma-related if it met one of the following criteria: (1) the encounter had a primary ICD-10 diagnosis code for asthma, or (2) the encounter included a maximum of two ICD-10 codes with at least one being an asthma diagnosis code. Encounters were limited to those from specialty departments most relevant to asthma care (i.e., Internal Medicine, Pulmonary, Family Practice, Emergency, Allergy/Immunology, Otorhinolaryngology, Gerontology, Obstetrics/Gynecology, and Hospital Medicine) that were present throughout all years of the 8-year study period. We also examined how trends in asthma healthcare utilization during 2020 varied by sex, age, race, and specialty care.

### Asthma drug prescriptions

Weekly counts of prescriptions for ICS, OCS and SABA were examined for each of the selected time periods. The ICS category included ICS-only formulations and ICS combination inhalers (i.e., with long-acting beta agonists [LABAs], long-acting muscarinic antagonists [LAMAs], and/or SABAs). Prescriptions were limited to those associated with a previously defined asthma-related encounter and were selected by name, dosage, and administration route to ensure they were for asthma-related conditions (Supplemental Table 1). If multiple medications of the same type were reported in an asthma-related encounter, only one medication from that medication type was counted.

### Time periods

Distinct weekly intervals in 2020 were identified to characterize the within-year changes in asthma healthcare utilization due to pandemic responses. In Philadelphia, COVID-19 restrictions began with business closures on March 16, 2020 (Week 12) followed by a stay-at-home order on March 23, 2020 (Week 13), with a phased reopening beginning June 5, 2020 (Week 23). We empirically identified Weeks 9 and 19 of 2020 as key inflection points for analyses based on inspection of weekly encounter patterns. These timepoints were selected to capture the transition periods when healthcare-seeking behaviors began changing in anticipation of, and in response to, pandemic restrictions. The pre-pandemic period encompassed the weekly encounter counts averaged across 2017-2019. The pandemic period included the weekly encounter counts for 2020. The post-pandemic period represented the weekly encounter counts averaged across 2021-2024. This approach allowed us to compare asthma healthcare utilization patterns before, during, and after the onset of the COVID-19 pandemic.

### Statistical methods

Demographic characteristics of unique patients with asthma-related encounters within each time period (pre-pandemic, pandemic, and post-pandemic) were summarized using standard descriptive statistics. To further evaluate whether associations between patient characteristics and healthcare encounters within each time period varied over time, we fitted generalized linear mixed-effects models with a logit link and random intercepts to account for repeated visits across periods. Each demographic or clinical predictor was a fixed effect along with its interaction with time period. Statistical significance was defined as p ≤ 0.001, after correcting for multiple comparisons across clinical and demographic predictors (0.05/36 comparisons) and type II Wald chi-square tests were used to assess interaction significance. Changes in weekly asthma-related healthcare encounters and drug prescriptions were evaluated by fitting quasi-Poisson generalized linear models that included time-period, modeled as a categorical variable (2017-2019, 2020, and 2021-2024), weekly intervals (Weeks 1-8, 9-18, 19-52) that captured major trend changes in 2020, year × weekly interval interaction terms and an offset to account for differences in length of pre-pandemic and post-pandemic periods. Deviance-based residuals were examined to assess model diagnostics and overall goodness of fit. Statistical significance was set at p ≤ 0.05. Analyses were conducted in R 4.3.2.

## RESULTS

### Patient characteristics

Over the course of the study period, a total of 42,242 unique individuals had a qualifying encounter (Supplemental Figure 1, Table 1). Included patients were primarily Female (69%), White (54%), non-Hispanic (95%), ≥35 years old (68%), Overweight/Obese (70%), Privately Insured (55%), and Never-Smokers (62%) compared to other levels of each corresponding variable. Generalized linear mixed models indicated that the associations between patient characteristics and having an encounter differed significantly across time periods (all p < 0.001 after Bonferroni correction). Changes included differences in age distributions, with patients in the 65+ years increasing in 2020, while those aged 18–24 decreased from 11% in 2017-2019 to 7.8% in 2020. The comorbidity burden of patients shifted beginning in 2020 and persisted post-pandemic: patients with an Elixhauser score of 0 decreased consecutively throughout all time periods from 82% in 2017-2019 to 72% in 2021-2024, while patients with Elixhauser scores <0, 1-9, and 10+ increased consecutively throughout all time periods. Data collection quality was impacted during 2020, with missing data increasing for both BMI and insurance status: Unknown BMI increased to 35% in 2020 versus 6.5% in 2017-2019 and 11% in 2021–2024, while Unknown insurance status increased to 18% in 2020 versus 5.2% pre-pandemic and 6.0% post-pandemic.

**Table 1:** Demographic and clinical characteristics of patients with asthma-related encounters by pre-pandemic (2017-2019), pandemic (2020), and post-pandemic (2021-2024) time periods (total unique n= 42,242).

| Characteristic | 2017-2019<br>N = 21,855* | 2020<br>N = 12,248* | 2021-2024<br>N = 27,469* | p-value<sup>‡</sup> | Overall<br>N = 42,242<sup>†</sup> |
| --- | --- | --- | --- | --- | --- |
| **Sex** |  |  |  | **<0.001** |  |
| Female | 15,000 (69%) | 8,665 (71%) | 19,130 (70%) |  | 29,032 (69%) |
| Male | 6,855 (31%) | 3,583 (29%) | 8,339 (30%) |  | 13,210 (31%) |
| **Race** |  |  |  | **<0.001** |  |
| AIAN* | 34 (0.2%) | 26 (0.2%) | 55 (0.2%) |  | 79 (0.2%) |
| Asian | 616 (2.8%) | 312 (2.5%) | 932 (3.4%) |  | 1,366 (3.2%) |
| Black | 8,067 (37%) | 4,582 (37%) | 10,503 (38%) |  | 15,705 (37%) |
| NHPI<sup>∞</sup> | 30 (0.1%) | 9 (<0.1%) | 51 (0.2%) |  | 76 (0.2%) |
| White | 12,061 (55%) | 6,703 (55%) | 14,426 (53%) |  | 22,814 (54%) |
| Other<sup>†</sup> | 1,047 (4.8%) | 616 (5.0%) | 1,502 (5.5%) |  | 2,202 (5.2%) |
| **Ethnicity** |  |  |  | **<0.001** |  |
| Hispanic/Latino | 703 (3.2%) | 434 (3.5%) | 1,193 (4.3%) |  | 1,641 (3.9%) |
| non-Hispanic/Latino | 21,034 (96%) | 11,721 (96%) | 26,024 (95%) |  | 40,269 (95%) |
| Unknown/Other | 118 (0.5%) | 93 (0.8%) | 252 (0.9%) |  | 332 (0.8%) |
| **Age** |  |  |  | **<0.001** |  |
| 18-24 | 2,351 (11%) | 953 (7.8%) | 2,612 (9.5%) |  | 5,293 (13%) |
| 25-34 | 3,927 (18%) | 2,256 (18%) | 5,077 (18%) |  | 8,251 (20%) |
| 35-44 | 3,747 (17%) | 2,131 (17%) | 4,767 (17%) |  | 7,277 (17%) |
| 45-54 | 3,983 (18%) | 2,183 (18%) | 4,501 (16%) |  | 7,110 (17%) |
| 55-64 | 4,069 (19%) | 2,265 (18%) | 4,864 (18%) |  | 7,292 (17%) |
| 65-74 | 2,668 (12%) | 1,724 (14%) | 3,827 (14%) |  | 4,963 (12%) |
| 75-84 | 946 (4.3%) | 634 (5.2%) | 1,554 (5.7%) |  | 1,763 (4.2%) |
| 85+ | 164 (0.8%) | 102 (0.8%) | 267 (1.0%) |  | 293 (0.7%) |
| **BMI** |  |  |  | **<0.001** |  |
| Not Overweight or Obese | 4,986 (23%) | 1,811 (15%) | 5,692 (21%) |  | 9,721 (23%) |
| Overweight | 5,719 (26%) | 2,128 (17%) | 6,795 (25%) |  | 11,120 (26%) |
| Obesity Class I | 4,342 (20%) | 1,747 (14%) | 5,403 (20%) |  | 8,473 (20%) |
| Obesity Class II | 2,599 (12%) | 1,092 (8.9%) | 3,201 (12%) |  | 5,018 (12%) |
| Obesity Class III | 2,779 (13%) | 1,123 (9.2%) | 3,249 (12%) |  | 5,122 (12%) |
| Unknown/Other | 1,430 (6.5%) | 4,347 (35%) | 3,129 (11%) |  | 2,788 (6.6%) |
| **Insurance** |  |  |  | **<0.001** |  |
| Medicaid | 3,764 (17%) | 1,804 (15%) | 5,127 (19%) |  | 7,886 (19%) |
| Medicare | 4,537 (21%) | 2,433 (20%) | 6,446 (23%) |  | 9,567 (23%) |
| Private | 12,426 (57%) | 5,769 (47%) | 14,237 (52%) |  | 23,421 (55%) |
| Unknown/Other | 1,128 (5.2%) | 2,242 (18%) | 1,659 (6.0%) |  | 1,368 (3.2%) |
| **Smoking status** |  |  |  |  |  |
| Current Smoker | 1,919 (8.8%) | 920 (7.5%) | 2,162 (7.9%) |  | 3,581 (8.5%) |
| Former Smoker<sup>§</sup> | 5,921 (27%) | 3,406 (28%) | 6,987 (25%) |  | 10,662 (25%) |
| Never Smoked | 13,369 (61%) | 7,703 (63%) | 17,287 (63%) |  | 26,233 (62%) |
| Unknown/Other | 646 (3.0%) | 219 (1.8%) | 1,033 (3.8%) |  | 1,766 (4.2%) |
| **Elixhauser** |  |  |  | **<0.001** |  |
| <0 | 590 (2.7%) | 423 (3.5%) | 1,104 (4.0%) |  | 1,495 (3.5%) |
| 0 | 17,999 (82%) | 9,535 (78%) | 19,641 (72%) |  | 31,384 (74%) |
| 1-9 | 2,508 (11%) | 1,703 (14%) | 4,665 (17%) |  | 6,505 (15%) |
| 10+ | 758 (3.5%) | 587 (4.8%) | 2,059 (7.5%) |  | 2,858 (6.8%) |

\*Total number of unique patients with healthcare encounters in each time period (%)
†Total number of unique patients included in analyses. Note that some patients had encounters across multiple time periods, so this is not the sum of the time period columns.
‡ Mixed-effects logistic regression models testing whether the association between each characteristic and healthcare utilization differs significantly across time periods, accounting for repeated observations. Bonferroni-corrected p-values ( $\alpha = 0.0014$ ) for multiple testing correction across demographic characteristics.
¶ Other race includes those that identify with a race not listed in the EHR or the "Multi" race category.
¥ AIAN = American Indian/Alaska Native
∞ NHPI = Native Hawaiian/Pacific Islander
§Former Smoker includes those with "Smoker, current status unknown" category in the EHR

### Asthma healthcare utilization trends

Prior to the implementation of public health measures in March 2020, asthma healthcare utilization remained similar to pre-pandemic trends observed in 2017-2019 (Figure 1). During the first eight weeks of 2020, weekly encounters averaged 429, comparable to historical 2017-2019 patterns. Utilization then surged to 604 weekly encounters during Weeks 9-18 before returning to pre-pandemic levels in Weeks 19-52 of 2020, and the following weeks in 2021-2024 (Supplemental Table 2).

**Figure 1.**
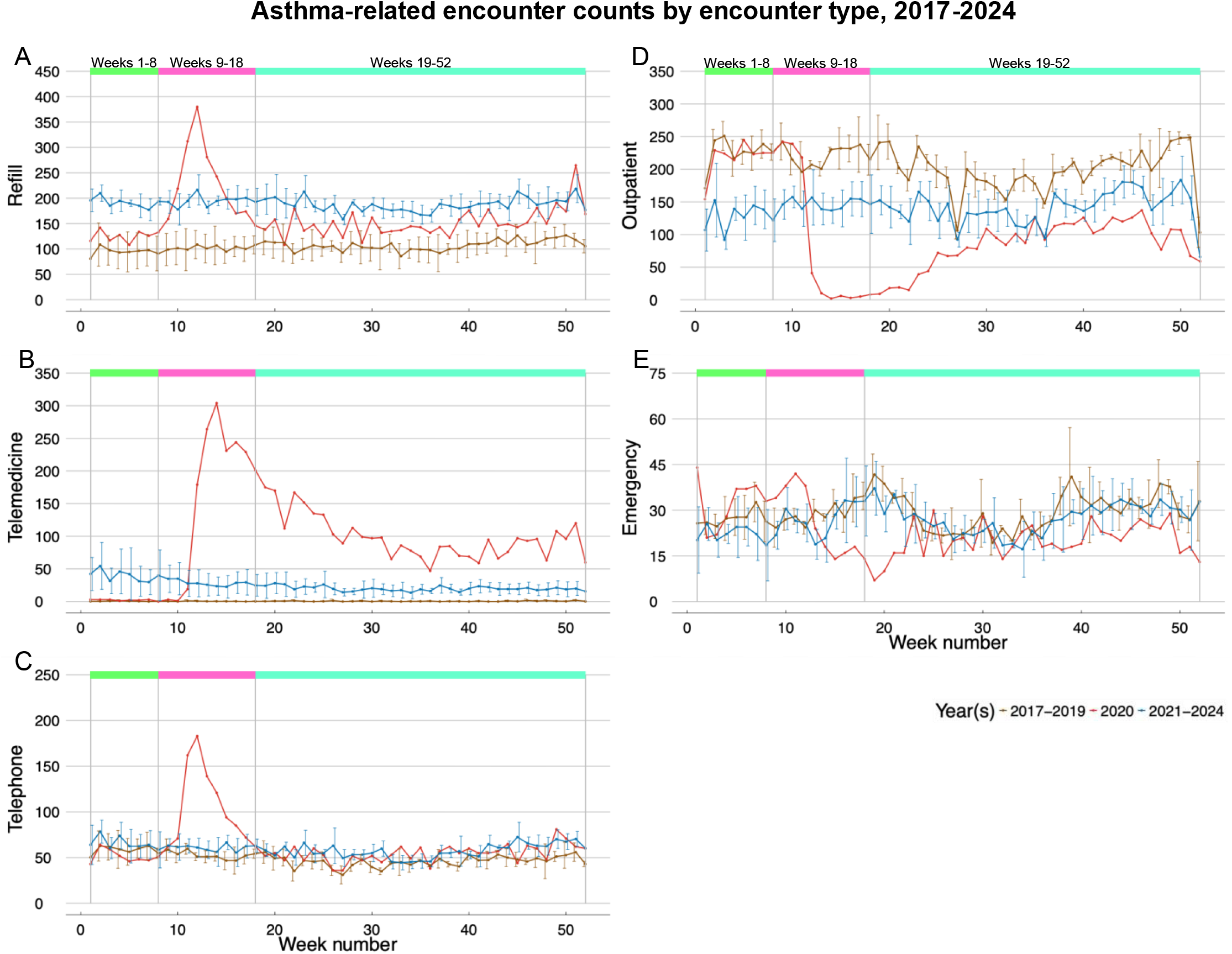
Trends in refill (1A), telemedicine (1B), telephone (1C), outpatient (1D), and emergency (1E) encounters, each shown in separate plots. The x-axis represents the week number relative to January 1, while the y-axis indicates the number of asthma-related encounters. Each plot features three lines: one for weekly average counts from 2017 to 2019 (brown), one for weekly counts in 2020 (red), and one for weekly average counts from 2021 to 2024 (blue). Additionally, separation lines and colored boxes mark relevant week intervals in 2020, Weeks 1-8, 9-18, and 19-52. Week 53 was omitted due to its shorter length.

Prescription refill patterns changed throughout the study period (Figure 1A). Comparing Weeks 1-8 and 9-18 of the pre-pandemic years to 2020, refill encounters increased 32%(RR=1.32, 95% CI: 1.10-1.58) and 123% (RR=2.23, 95% CI: 1.96-2.55), respectively (Table 2). While counts subsided during Weeks 19-52 of 2020, refills remained 42% above the corresponding pre-pandemic weeks (RR=1.42, 95% CI: 1.31-1.54). Increased refills persisted through 2021-2024, doubling pre-pandemic rates in Weeks 1-8 (RR=2.02, 95% CI: 1.79-2.27) and continuing near these levels in Weeks 9-18 (RR=1.90, 95% CI: 1.72-2.11) and Weeks 19-52 (RR=1.74, 95% CI: 1.65-1.84). Telemedicine encounters had the largest change (Figure 1B). While negligible in 2017-2019, telemedicine surged in 2020: 36,445% higher during Weeks 9-18 (RR=365.45, 95% CI: 46.68-2860.95) and 13,869% higher during Weeks 19-52 (RR=138.69, 95% CI: 55.01-349.69) compared to pre-pandemic. Telemedicine remained integrated into asthma care during 2021-2024: 7,465% higher in Weeks 1-8 (RR=75.65, 95% CI: 9.64-593.48), 6,015% higher in Weeks 9-18 (RR=61.15,

**Table 2:** Rate ratios (RRs) for adult asthma-related encounters by time periods and weekly intervals considered. Models compared weekly intervals of 2020 and 2021-2014 to corresponding weekly intervals in pre-pandemic years (2017–2019). Encounter counts for each outcome (Refill, Telemedicine, Telephone, Outpatient, and Emergency) across relevant weekly intervals are presented as RR (95% CI) from generalized linear regression models. Combined effects were calculated using estimated marginal means with Dunnett adjustment for multiple comparisons (critical p-value = 0.0085, α = 0.05) using the pre-pandemic (2017-2019) weekly intervals as reference.

| Encounter Type<br>(N) | Weeks 1–8<br>RR (95% CI) | p-value | Weeks 9–18<br>RR (95% CI) | p-value | Weeks 19–52<br>RR (95% CI) | p-value |
| --- | --- | --- | --- | --- | --- | --- |
| **Refill**<br>**(64,138)** |  |  |  |  |  |  |
| 2017–2019 | 1.00 (ref) |  | 1.00 (ref) |  | 1.00 (ref) |  |
| 2020 | 1.32 (1.10–1.58) | 0.001 | 2.23 (1.96–2.55) | <0.001 | 1.42 (1.31–1.54) | <0.001 |
| 2021–2024 | 2.02 (1.79–2.27) | <0.001 | 1.90 (1.72–2.11) | <0.001 | 1.74 (1.65–1.84) | <0.001 |
| **Telemedicine**<br>**(10,175)** |  |  |  |  |  |  |
| 2017–2019 | 1.00 (ref) |  | 1.00 (ref) |  | 1.00 (ref) |  |
| 2020 | 4.64 (0.33–64.47) | 0.333 | 365.45 (46.68–2860.95) | <0.001 | 138.69 (55.01–349.69) | <0.001 |
| 2021–2024 | 75.65 (9.64–593.48) | <0.001 | 61.15 (7.78–480.26) | <0.001 | 28.07 (11.11–70.90) | <0.001 |
| **Telephone**<br>**(23,306)** |  |  |  |  |  |  |
| 2017–2019 | 1.00 (ref) |  | 1.00 (ref) |  | 1.00 (ref) |  |
| 2020 | 0.88 (0.70–1.10) | 0.338 | 2.00 (1.70–2.35) | <0.001 | 1.20 (1.07–1.34) | 0.001 |
| 2021–2024 | 1.12 (0.98–1.29) | 0.120 | 1.16 (1.02–1.32) | 0.020 | 1.25 (1.16–1.35) | <0.001 |
| **Outpatient**<br>**(66,729)** |  |  |  |  |  |  |
| 2017–2019 | 1.00 (ref) |  | 1.00 (ref) |  | 1.00 (ref) |  |
| 2020 | 0.97 (0.72–1.29) | 0.939 | 0.35 (0.24–0.52) | <0.001 | 0.43 (0.35–0.54) | <0.001 |
| 2021–2024 | 0.57 (0.46–0.71) | <0.001 | 0.67 (0.55–0.80) | <0.001 | 0.71 (0.64–0.79) | <0.001 |
| **Emergency**<br>**(11,259)** |  |  |  |  |  |  |
| 2017–2019 | 1.00 (ref) |  | 1.00 (ref) |  | 1.00 (ref) |  |
| 2020 | 1.19 (0.91–1.57) | 0.271 | 0.88 (0.68–1.15) | 0.481 | 0.68 (0.58–0.80) | <0.001 |
| 2021–2024 | 0.82 (0.67–1.00) | 0.051 | 0.94 (0.79–1.11) | 0.597 | 0.92 (0.84–1.01) | 0.110 |

95% CI: 7.78-480.26), and 2,707% higher in Weeks 19-52 (RR=28.07, 95% CI: 11.11-70.90) versus 2017-2019. Telephone encounters were 100% higher during Weeks 9-18 of 2020 (RR=2.00, 95% CI: 1.70-2.35) compared to the same weeks in pre-pandemic. However, unlike other encounter types, telephone encounters returned to pre-pandemic levels in Weeks 1-8 and Weeks 9-18 post-pandemic but remained slightly elevated in Weeks 19-52 of 2020 and 2021-2024 (2020: RR=1.20, 95% CI: 1.07-1.34; 2021-2024: RR=1.25, 95% CI: 1.16-1.35) (Figure 1C). In contrast to the expansion of refill and telemedicine care options, outpatient encounters experienced significant and persistent disruptions. Outpatient encounters declined precipitously during 2020, with 65% fewer encounters in Weeks 9-18 (RR=0.35, 95%: CI 0.24-0.52) and 57% fewer encounters in Weeks 19-52 (RR=0.43, 95%: CI 0.35-0.54) compared to their respective pre-pandemic weeks. These reductions persisted into the post-pandemic period, with outpatient encounters remaining 29-43% lower than pre-pandemic levels across all time intervals during 2021-2024 (Figure 1D). Emergency encounters during Weeks 1-8 and 9-18 of 2020 remained at pre-pandemic levels, but were 32% lower during in Weeks 19-52 compared to the respective weekly pre-pandemic time period (RR=0.68, 95% CI: 0.58-0.80). In 2021-2024, emergency encounters returned to their pre-pandemic levels (Figure 1E).

### Asthma drug trends

Prior to COVID-19–related public health measures in March 2020, adult asthma prescription patterns at Penn Medicine in Weeks 1-8 were similar to those during the same weeks of 2017-2019 (Figure 2). In 2020, however, prescribing patterns changed for all medication classes, with temporal patterns emerging for maintenance therapy (SABA and ICS) versus exacerbation therapy (OCS) (Table 3). SABA prescriptions surged dramatically during Weeks 9-18 of 2020, with a 73% increase compared to Weeks 9-18 in 2017-2019 (RR=1.73, 95% CI: 1.50-2.00) (Figure 2A). While this surge subsided during Weeks 19-52 of 2020 and returned to pre-pandemic levels (RR=1.04, 95% CI: 0.94-1.14), SABA prescription rates remained modestly elevated throughout 2021-2024, with rates 19-21% higher than pre-pandemic levels during Weeks 9-18 and Weeks 19-52 (Weeks 9-18: RR=1.21, 95% CI: 1.08-1.35; Weeks 19-52: RR=1.19, 95% CI: 1.12-1.27). During Weeks 9-18 of 2020, ICS prescriptions showed a similar pattern to SABA, with a significant 43% increase compared to Weeks 9-18 in 2017-2019 (RR=1.43, 95% CI: 1.23-1.66). Like SABA, ICS prescriptions returned to pre-pandemic levels during Weeks 19-52 of 2020 but had a statistically significant increase of 15% in Weeks 19-52 post-pandemic (Figure 2B). OCS prescriptions showed a pattern opposite to that of SABA and ICS, characterized by sustained reductions at the outset of the pandemic. Weeks 9-18 of 2020 had a non-statistically significant decline of OCS prescriptions by 5% (RR=0.95, 95% CI: 0.76-1.19) compared to Weeks 9-18 in 2017-2019, whereas Weeks 19-52 of 2020 declined by 37% compared to the same weekly interval in 2017-2019 (RR=0.63, 95% CI: 0.55-0.74). This reduction was sustained across all weekly intervals in 2021-2024, with 22-42% fewer OCS prescriptions (Weeks 1-8: RR=0.58, 95% CI: 0.49-0.69; Weeks 9-18: RR=0.70, 95% CI: 0.59-0.81; Weeks 19-52: RR=0.78, 95% CI: 0.72-0.85) (Figure 2C).

**Table 3:** Models compare week intervals in 2020 and 2021–2024 to pre-pandemic week intervals (2017–2019). Encounter counts for each visit with an asthma-related medication (SABA, ICS, and OCS) across relevant week intervals are presented as RR (95% CI) using quasi-Poisson generalized linear regression. Combined effects were calculated using estimated marginal means. Statistical significance was assessed using Dunnett’s method to adjust for multiple comparisons, with a critical p-value of 0.0085 (α = 0.05, 6 comparisons per medication type).

| **Encounter Medication (N)** | **Weeks 1–8 RR (95% CI)** | **p-value** | **Weeks 9–18 RR (95% CI)** | **p-value** | **Weeks 19–52 RR (95% CI)** | **p-value** |
| --- | --- | --- | --- | --- | --- | --- |
| **SABA (71,161)** |  |  |  |  |  |  |
| **2017–2019** | 1.00 (ref) |  | 1.00 (ref) |  | 1.00 (ref) |  |
| **2020** | 1.11 (0.92–1.34) | 0.337 | 1.73 (1.50–2.00) | <0.001 | 1.04 (0.94–1.14) | 0.570 |
| **2021–2024** | 1.13 (1.00–1.28) | 0.047 | 1.21 (1.08–1.35) | <0.001 | 1.19 (1.12–1.27) | <0.001 |
| **ICS (59,037)** |  |  |  |  |  |  |
| **2017–2019** | 1.00 (ref) |  | 1.00 (ref) |  | 1.00 (ref) |  |
| **2020** | 1.10 (0.91–1.32) | 0.427 | 1.43 (1.23–1.66) | <0.001 | 1.06 (0.96–1.16) | 0.341 |
| **2021–2024** | 1.13 (1.00–1.28) | 0.042 | 1.15 (1.03–1.28) | 0.009 | 1.15 (1.08–1.22) | <0.001 |
| **OCS (25,787)** |  |  |  |  |  |  |
| **2017–2019** | 1.00 (ref) |  | 1.00 (ref) |  | 1.00 (ref) |  |
| **2020** | 0.93 (0.73–1.18) | 0.714 | 0.95 (0.76–1.19) | 0.821 | 0.63 (0.55–0.74) | <0.001 |
| **2021–2024** | 0.58 (0.49–0.69) | <0.001 | 0.70 (0.59–0.81) | <0.001 | 0.78 (0.72–0.85) | <0.001 |

**Figure 2.**
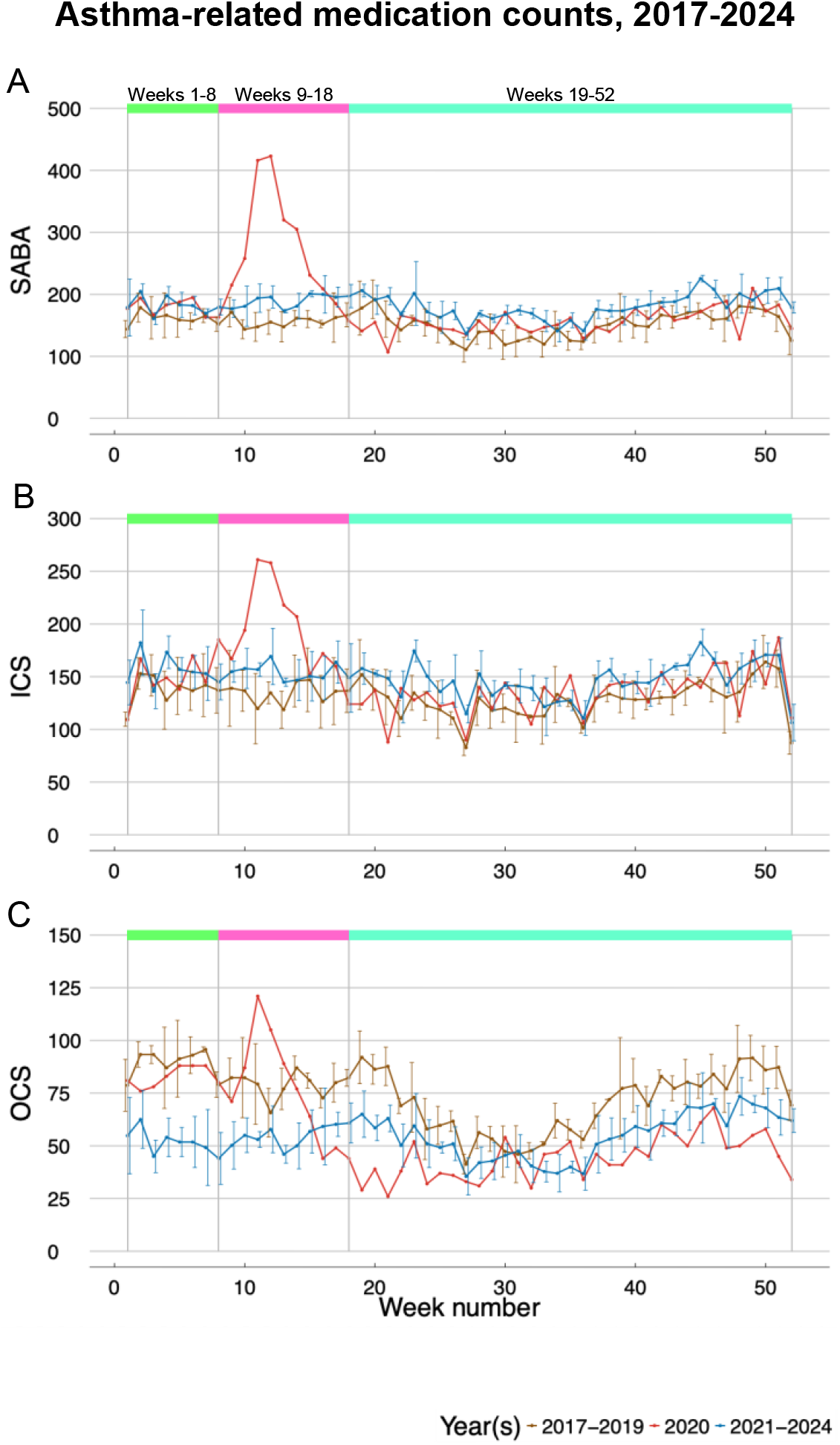
Trends in asthma-related encounters where SABA (2A), ICS (2B), and OCS (2C) medications were prescribed, each shown in separate plots. The x-axis represents the week number, while the y-axis indicates the number of asthma-related encounters and with an asthma-related medication. Each plot features three lines: one for weekly average counts from 2017 to 2019 (brown), one for weekly counts in 2020 (red), and one for weekly average counts from 2021 to 2024 (blue). Additionally, separation lines and colored boxes mark relevant week intervals in 2020, Weeks 1-8, 9-18, and 19-52. Week 53 was omitted due to its shorter length.

### Differences in trends by sex, age, race, and specialty care

Sex-, age-, race-, and specialty care-specific encounter trends in 2020 are presented in Figure 3. Encounters showed similar surge patterns during Weeks 9-18 of 2020 across most demographic groups, corresponding to the initial weeks of the first lockdown in the Greater Philadelphia region. Female and Male patients exhibited similar trends with peak encounters during Week 12 of 2020 (Figure 3A), however, Female patients had a sharper increase in encounters compared to Male patients. The 25-34, 35-44, 45-54, and 55-64 age groups had the sharpest increases in encounters during the initial pandemic period (i.e., Weeks 9-18 versus Weeks 1-8 of 2020), while those in the 18-24, 65-74, and 75-84 age categories showed slight increases, and those aged 85+ had no changes throughout 2020. Encounters in all age groups returned to pre-pandemic levels by Week 18 (Figure 3B).

**Figure 3.**
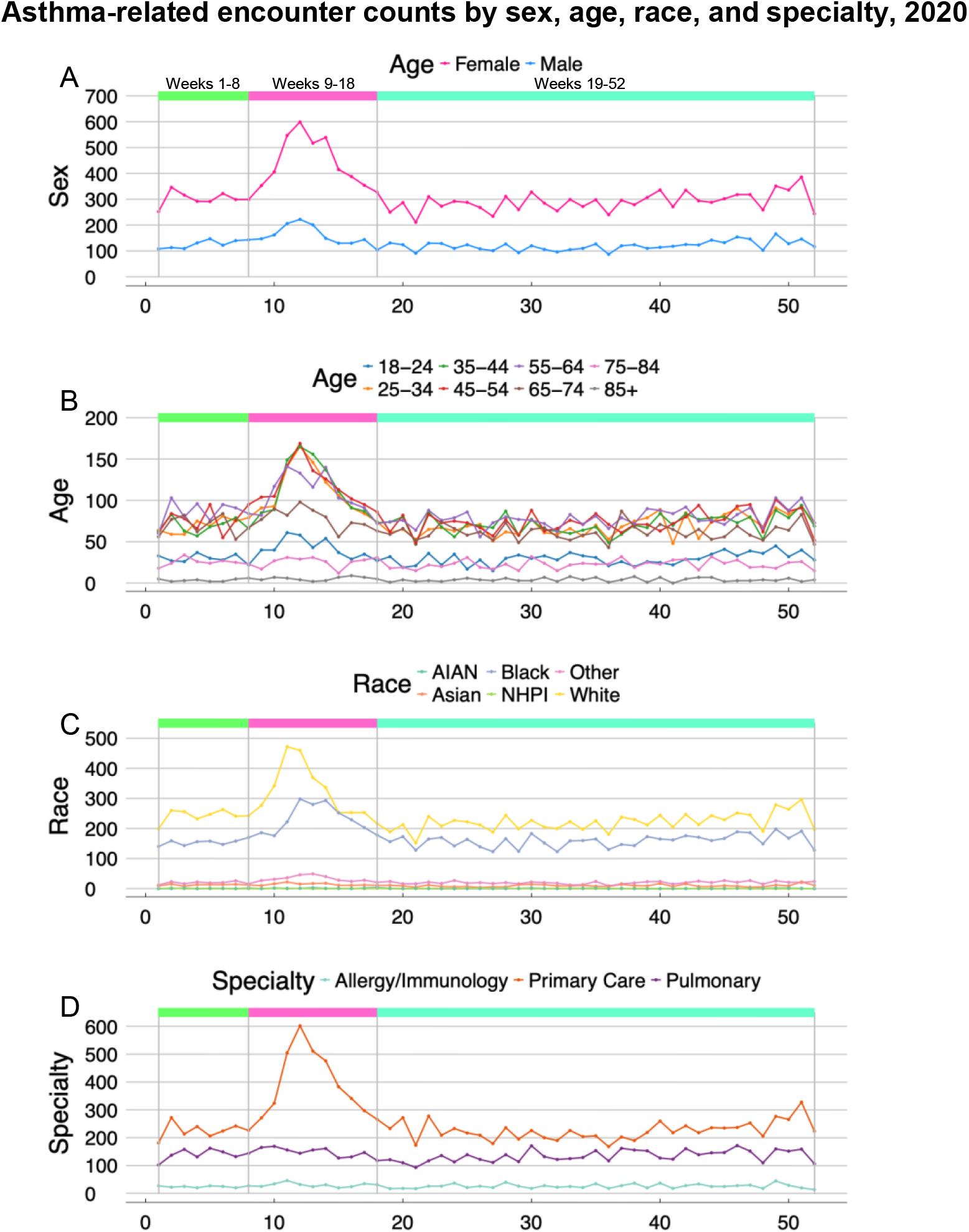
Weekly counts of asthma-related encounters by sex, age group, race, and specialty care in 2020. The x-axis represents the week number, while the y-axis indicates the number by sex, age group, race, and specialty care in 2020. Separation lines and colored boxes mark relevant week intervals in 2020, Weeks 1-8, 9-18, and 19-52. Week 53 was omitted due to its shorter length. Panel A: Weekly counts of asthma-related encounters in 2020 by sex (Female and Male). Panel B: Weekly counts of asthma-related encounters in 2020 by age groups (18-24, 25-34, 35-44, 45-54, 55-64, 65-74, 75-84, and 85+). Panel C: Weekly counts of asthma-related encountersin 2020 by race group (American Indian and Alaska Native (AIAN), Asian, Black, Native Hawaiian and Pacific Islander (NHPI), White, and Unknown/Other). Panel D: Weekly counts of asthma-related encounters in 2020 by specialty care (Allergy/Immunology, Primary Care, and Pulmonary). Primary Care is defined as Family Practice, Internal Medicine, and Gerontology specialty departments.

In terms of race, the sharpest increases in encounters during Weeks 9-18 of 2020 were observed for White and Black patients, the categories representing 91% of unique patients. Asian and Other patients showed small increases in encounter counts. Native Hawaiian/Pacific Islander (NHPI) and American Indian/Alaska Native (AIAN) patients had fewer encounters overall and showed no changes throughout 2020. The weekly increase in encounters during the onset of the pandemic occurred earlier for White patients than those of other race categories (Figure 3C). For encounter counts by specialty care, Primary Care departments defined by Family Practice, Internal Medicine, and Gerontology departments, increased in Weeks 9-18 of 2020 whereas Allergy/Immunology and Pulmonary departments did not show changes throughout 2020 (Figure 3D).

related medication prescription trends showed similar patterns across sex, age, and race categories, mirroring the encounter trends observed in Figure 3. Thus, the observed increases in healthcare utilization and medication prescribing during the early pandemic period were not driven by patients of these specific demographic subgroups. Demographic characteristics for patients with encounters and prescriptions during week intervals in 2020 are presented in Supplemental Tables 3 and 4. Primary Care encounters in 2020 also showed similar patterns to medication prescription trends, indicating that primary care encounters drove the increase in medication prescriptions in Weeks 9-18 of 2020 compared to Allergy/Immunology and Pulmonary departments.

## DISCUSSION

This retrospective analysis of 2017-2024 data from a multi-hospital EHR database in greater Philadelphia found that shortly after the COVID-19 lockdown began (Weeks 9-18 of 2020) refill, telemedicine, and telephone encounters increased 123%, 36,445%, and 100%, respectively, while outpatient encounters decreased 65% compared to 2017-2019. Emergency encounters decreased 12% during lockdown and 32% during Weeks 19-52 of 2020 versus pre-pandemic periods. These findings align with population-based studies from Scotland and Wales that documented a 36% reduction in emergency asthma admissions during national lockdown periods in pediatric and adult populations^15^ as well as reports from Kaiser Permanente Colorado describing the transition from in-person to virtual asthma care during the COVID-19 pandemic in adults.^4^

Our analyses of medication prescriptions revealed maintenance therapy drugs SABA and ICS surged by 73% and 43%, respectively, during Weeks 9-18 of 2020, compared to Weeks 9-18 in pre-pandemic years. Our medication prescription findings partially align with a UK study that reported a 121% spike in ICS prescriptions preceding lockdown implementation in pediatric and adult populations.^12^ However, that study observed increased OCS prescriptions during lockdown. This divergence in exacerbation therapy drug patterns may reflect differences in healthcare systems, patient populations, prescribing practices for maintenance therapy in severe asthma versus acute exacerbation management, or variations in medication stockpiling behaviors.

Unlike children who showed decreased prescriptions at lockdown onset, ^13–15^ adults in our study increased maintenance medications while decreasing OCS, suggesting proactive stockpiling. The persistence of increased SABA and ICS use through 2024 indicates longer-term adaptations. Whether these changes reflect patient preferences, provider practices, new guidelines (GINA 2021),^16^, or continued effects of the pandemic on healthcare-seeking behavior is beyond the scope of the current study.

Temporal trends stratified by sex, race, age, and primary care demonstrated consistency in utilization during 2020, with counts proportional to patient percentages in each stratum. Female patients had higher overall utilization and sharper increase in healthcare encounters during lockdown weeks compared to Male patients, consistent with known epidemiologic asthma prevalence patterns in the general population. While all racial groups exhibited sharp increases at the pandemic’s onset, distinct temporal patterns emerged in peak utilization timing. White patients had an earlier surge during Weeks 9-10 of 2020, coinciding with pandemic awareness that preceded the lockdown. Black patients, who comprised 37% of our study population, had encounter peaks beginning at Week 11, suggesting a one-week temporal delay that may reflect documented disparities in healthcare access, pandemic information dissemination, or differential levels of healthcare system engagement observed in studies of pandemic-related healthcare utilization.^17,18^ Age-specific encounter trends during 2020 showed the youngest (18-24 years) and oldest (85+ years) adults maintained fewer encounters throughout the year with no change during pandemic onset, while adults in the 65-84 year range had a proportionately smaller increase in encounters during the pandemic onset compared with the steeper rise among 25-64 year-olds. These patterns may reflect the intersection of several factors: stricter self-isolation behaviors in response to public health recommendations were possible among the youngest and oldest adults, and technological barriers to accessing telemedicine services and possible concerns about COVID-19 exposure risk in healthcare settings among older adults.^19^ Further, we found primary care contributed the most to the increase in healthcare utilization during Weeks 9-18 in 2020. In comparison, Allergy/Immunology and Pulmonary departments remained stable even after the lockdown onset. These specialties manage fewer patients but with more specialized asthma care overall, compared to primary care providers. This pattern is consistent with reports of a US-Canadian expert panel^20^ of allergy/immunology specialists which emphasized prioritizing severe asthma cases for continued in-person allergy care, while postponing of face-to-face interactions.

Several limitations may affect the interpretation of our findings. Our analysis was restricted to a single health system in Philadelphia, which may limit generalizability to other geographic areas or healthcare systems with different patient populations, resources, or pandemic response strategies. For instance, the health system’s established telehealth infrastructure may have facilitated the rapid transition to remote care more effectively than systems without such capabilities. There was a 28.5% increase in patients with unknown BMI from 2017-2019 to 2020. This shift likely reflects virtual care encounters and associated limitations in data collection. These data quality issue findings have been reported in other telemedicine studies during COVID-19, with up to one-third of patients missing race, ethnicity, or language data^17–19^. Generalizability to patients with more severe asthma who would have required hospitalization or specialized treatments is limited as we excluded inpatient encounters and biologic prescriptions from formal analyses due to low counts. Further, we included clinical sites that were in practice for all 8 years of the study, and thus we did not capture healthcare utilization in newer sites. Although we explored demographic trends, our analyses did not incorporate socioeconomic variables or other social determinants of health that could further explain shifts in care-seeking behavior. Lastly, we did not have data about deaths that occurred in the system during the study period, and therefore, there is a potential for survival bias that would arise from depletion of patients with worse asthma conditions.

Our study strengths include the study design and comprehensive EHR dataset spanning 8 years that captured healthcare utilization patterns in a major U.S. metropolitan area. The follow-up through 2024 allowed us to distinguish between temporary disruptions and lasting transformations in healthcare delivery, which is not available in studies with shorter observation periods. Reports of healthcare utilization changes due to the COVID-19 pandemic have primarily focused on pediatric populations; however, we included a large sample of adults with asthma to address an important research gap.

## CONCLUSION

The COVID-19 pandemic reshaped patterns of asthma care among adults, prompting reliance on telemedicine and medication refills with concomitant reductions in asthma outpatient and emergency encounters. Although these changes likely helped sustain asthma care during 2020, they also introduced new complexities, including declines in outpatient visits that persisted in post-pandemic years, especially among Black patients and those 65-84 years of age. Future research should investigate the long-term implications of these shifts for asthma outcomes and anticipate similar changes in healthcare-seeking behaviors during any future pandemic events.

## Data Availability

Data was not collected with informed consent, and based on ethical and legal considerations, the electronic health record (EHR) data used in this work cannot be shared widely. Upon reasonable request to the authors, data may be provided.

## Abbreviations/Acronyms

OCS: Oral corticosteroids
ICS: inhaled corticosteroids
EHR: Electronic health records
SABA: short-acting beta agonist
LABA: long-acting beta agonist
LAMA: long-acting muscarinic antagonist
COPD: chronic obstructive pulmonary disease
IRR: incidence rate ratios.

