## Supplemental Data for "Impact of the COVID-19 Pandemic on Adult Asthma-Related Healthcare Utilization"

**Supplemental Figures**

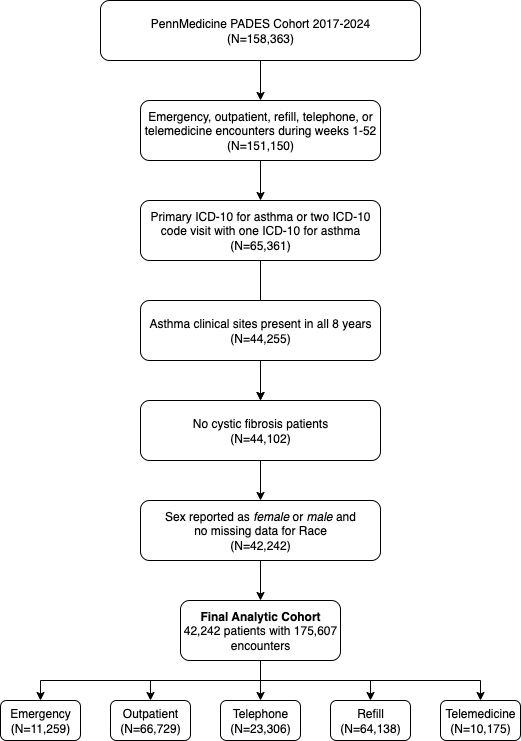

Figure E 1: Flowchart of patient selection for encounter analyses. Overview of steps followed to select final set of patients (N=42,242) from Penn Medicine EHR data spanning 2017-2024 and corresponding to 158,363 total encounters.

**Supplemental Tables**

Table E1: Asthma-related medications by type. Generic and brand medication names as recorded in the EHR.

| Medication Type | Medication Name |
| --- | --- |
| SABA | Albuterol, Albuterol Sulfate, Levalbuterol HCL, Levalbuterol Tartrate, Pirbuterol Acetate, Proair Digihaler, Proair HFA, Proair Respiclick, Proventil HFA, Ventolin HFA, Xopenex, Xopenex HFA |
| ICS | Alvesco, Armonair Digihaler, Arnuity Ellipta, Asmanex, Asmanex HFA, Beclomethasone Dipropionate, Beclomethasone Dipropionate HFA, Budesonide, Ciclesonide, Flovent Diskus, Flovent HFA, Flunisolide HFA, Fluticasone Furoate, Fluticasone Propionate, Fluticasone Propionate Diskus, Fluticasone Propionate HFA, Mometasone Furoate, Pulmicort, Pulmicort Flexhaler, QVAR, QVAR Redihaler |
| ICS/LABA | Advair Diskus, Advair HFA, AirDuo Digihaler, AirDuo Respiclick, Breo Ellipta, Breyna, Budesonide-Formoterol Fumarate, Dulera, Fluticasone Furoate-vilanterol, Fluticasone-Salmeterol, Mometasone Furo-Formoterol, Symbicort, Wixela Inhub |
| ICS/LABA/LAMA | Breztri Aerosphere, Budeson-Glycopyrrol-formoterol, fluticasone-umeclidin-vilant, Trelegy Ellipta |
| ICS/SABA | Airsupra, Albuterol-Budesonide |
| OCS | Cortef, Dexamethasone, Hydrocortisone, Medrol, Methylprednisolone, Prednisolone, Prednisolone Sodium Phosphate, Prednisone |

Table E2: Average weekly encounter counts by year and week intervals. The average weekly encounters of refill, telemedicine, telephone, outpatient, and emergency encounters grouped by pivotal week intervals in 2020, Weeks 1-8, 9-18, and 19-52 and by each year of the study period.

|  | Week Intervals | | |
| --- | --- | --- | --- |
|  | Weeks 1-8 | Weeks 9-18 | Weeks 19-52 |
| 2017 | 351 | 355 | 347 |
| 2018 | 433 | 427 | 392 |
| 2019 | 434 | 432 | 403 |
| 2020 | 429 | 604 | 411 |
| 2021 | 429 | 464 | 437 |
| 2022 | 433 | 422 | 426 |
| 2023 | 456 | 479 | 440 |
| 2024 | 468 | 468 | 426 |

Table E3: Age, sex, and race characteristics of patients from 2020 asthma-related encounters by encounter types and week intervals. Demographic characteristics are reported per encounter, including all encounters from patients with multiple encounters within a week interval.

| **Characteristic** | **Refill** | | | **Telephone** | | | **Telemedicine** | | | **Outpatient** | | | **Emergency** | | |
| --- | --- | --- | --- | --- | --- | --- | --- | --- | --- | --- | --- | --- | --- | --- | --- |
|  | **Weeks**  **1-8 N = 1,004***^1^* | **Weeks**  **9-18 N = 2,284***^1^* | **Weeks**  **19-52 N = 5,181***^1^* | **Weeks**  **1-8 N = 410***^1^* | **Weeks**  **9-18**  **N = 1,051***^1^* | **Weeks**  **19-52 N = 1,862***^1^* | **Weeks**  **1-8 N = 17***^1^* | **Weeks**  **9-18 N = 1,675***^1^* | **Weeks**  **19-52 N = 3,325***^1^* | **Weeks**  **1-8 N = 1,739***^1^* | **Weeks**  **9-18 N = 774***^1^* | **Weeks**  **19-52 N = 2,916***^1^* | **Weeks**  **1-8 N = 260***^1^* | **Weeks**  **9-18 N = 256***^1^* | **Weeks**  **19-52 N = 684***^1^* |
| **Age** |  |  |  |  |  |  |  |  |  |  |  |  |  |  |  |
| 18-24 | 61  (6.1%) | 160  (7.0%) | 354  (6.8%) | 27  (6.6%) | 64  (6.1%) | 98  (5.3%) | 1  (5.9%) | 103  (6.1%) | 210  (6.3%) | 97  (5.6%) | 55  (7.1%) | 197  (6.8%) | 52  (20%) | 42  (16%) | 149  (22%) |
| 25-34 | 174 (17%) | 424  (19%) | 952  (18%) | 65  (16%) | 208 (20%) | 289  (16%) | 8  (47%) | 297  (18%) | 584  (18%) | 238  (14%) | 115 (15%) | 328  (11%) | 76  (29%) | 69  (27%) | 208  (30%) |
| 35-44 | 186 (19%) | 469  (21%) | 961  (19%) | 54  (13%) | 195 (19%) | 329  (18%) | 5  (29%) | 312  (19%) | 632  (19%) | 259  (15%) | 107 (14%) | 357  (12%) | 47  (18%) | 60 (  23%) | 136  (20%) |
| 45-54 | 197 (20%) | 482 (  21%) | 1,024 (20%) | 72  (18%) | 213 (20%) | 319  (17%) | 2  (12%) | 300  (18%) | 601  (18%) | 298  (17%) | 134 (17%) | 491  (17%) | 41  (16%) | 49  (19%) | 83  (12%) |
| 55-64 | 205 (20%) | 403 (  18%) | 1,001 (19%) | 83  (20%) | 197 (19%) | 384  (21%) | 1  (5.9%) | 307  (18%) | 590  (18%) | 365  (21%) | 160 (21%) | 630  (22%) | 31  (12%) | 25  (9.8%) | 70  (10%) |
| 65-74 | 130 (13%) | 258  (11%) | 639  (12%) | 79  (19%) | 132 (13%) | 329  (18%) | 0  (0%) | 244  (15%) | 502  (15%) | 335  (19%) | 136 (18%) | 587  (20%) | 11 (4.2%) | 8  (3.1%) | 33  (4.8%) |
| 75-84 | 45 (4.5%) | 74  (3.2%) | 211  (4.1%) | 21 (5.1%) | 36  (3.4%) | 91  (4.9%) | 0  (0%) | 94  (5.6%) | 179  (5.4%) | 133  (7.6%) | 51  (6.6%) | 279 (9.6%) | 2  (0.8%) | 3  (1.2%) | 5  (0.7%) |
| 85+ | 6  (0.6%) | 14  (0.6%) | 39  (0.8%) | 9  (2.2%) | 6  (0.6%) | 23  (1.2%) | 0  (0%) | 18  (1.1%) | 27  (0.8%) | 14  (0.8%) | 16  (2.1%) | 47  (1.6%) | 0  (0%) | 0  (0%) | 0  (0%) |
| **Sex** |  |  |  |  |  |  |  |  |  |  |  |  |  |  |  |
| Female | 694 (69%) | 1,639 (72%) | 3,520 (68%) | 299 (73%) | 807 (77%) | 1,373 (74%) | 15  (88%) | 1,296 (77%) | 2,503 (75%) | 1,253 (72%) | 548 (71%) | 2,091 (72%) | 156 (60%) | 155 (61%) | 394  (58%) |
| Male | 310 (31%) | 645  (28%) | 1,661 (32%) | 111 (27%) | 244 (23%) | 489  (26%) | 2  (12%) | 379  (23%) | 822  (25%) | 486  (28%) | 226 (29%) | 825  (28%) | 104 (40%) | 101 (39%) | 290  (42%) |
| **Race** |  |  |  |  |  |  |  |  |  |  |  |  |  |  |  |
| AIAN | 1 (<0.1%) | 5  (0.2%) | 6  (0.1%) | 0  (0%) | 2  (0.2%) | 2  (0.1%) | 0  (0%) | 7  (0.4%) | 3  (<0.1%) | 5  (0.3%) | 1  (0.1%) | 10  (0.3%) | 0  (0%) | 0  (0%) | 0  (0%) |
| Asian | 27 (2.7%) | 53  (2.3%) | 96  (1.9%) | 11 (2.7%) | 25  (2.4%) | 45  (2.4%) | 0  (0%) | 40  (2.4%) | 87  (2.6%) | 56  (3.2%) | 25  (3.2%) | 89  (3.1%) | 3  (1.2%) | 0  (0%) | 10  (1.5%) |
| Black | 364 (36%) | 797 (35%) | 2,018 (39%) | 150 (37%) | 431 (41%) | 774  (42%) | 9  (53%) | 683 (41%) | 1,276 (38%) | 508 (29%) | 221 (29%) | 772  (26%) | 200 (77%) | 185 (72%) | 555  (81%) |
| NHPI | 0  (0%) | 0  (0%) | 2  (<0.1%) | 1  (0.2%) | 1  (<0.1%) | 6  (0.3%) | 0  (0%) | 2  (0.1%) | 3  (<0.1%) | 0  (0%) | 0  (0%) | 0  (0%) | 1  (0.4%) | 1  (0.4%) | 0  (0%) |
| White | 558 (56%) | 1,320 (58%) | 2,780 (54%) | 232 (57%) | 526 (50%) | 964  (52%) | 7  (41%) | 848 (51%) | 1,773 (53%) | 1,100 (63%) | 486 (63%) | 1,931 (66%) | 43  (17%) | 51  (20%) | 91  (13%) |
| Other | 54 (5.4%) | 109 (4.8%) | 279  (5.4%) | 16 (3.9%) | 66  (6.3%) | 71  (3.8%) | 1  (5.9%) | 95  (5.7%) | 183  (5.5%) | 70  (4.0%) | 41  (5.3%) | 114 (3.9%) | 13 (5.0%) | 19  (7.4%) | 28  (4.1%) |
| *^1^* n (%) | | | | | | | | | | | | | | | |

Table E 4 Age, sex, and race characteristics of patients prescribed asthma related medication in 2020 by week intervals. Demographic characteristics are reported per encounter, including all encounters from patients with multiple encounters within a week interval.

| **Characteristic** | **SABA** | | | **ICS** | | | **OCS** | | |
| --- | --- | --- | --- | --- | --- | --- | --- | --- | --- |
|  | **Weeks**  **1-8 N = 1,430***^1^* | **Weeks**  **9-18 N = 2,719***^1^* | **Weeks**  **19-52 N = 5,308***^1^* | **Weeks**  **1-8 N = 1,205***^1^* | **Weeks**  **9-18 N = 1,913***^1^* | **Weeks**  **19-52 N = 4,568***^1^* | **Weeks**  **1-8 N = 662***^1^* | **Weeks**  **9-18 N = 751***^1^* | **Weeks**  **19-52 N = 1,504***^1^* |
| **Age** |  |  |  |  |  |  |  |  |  |
| 18-24 | 138 (9.7%) | 221 (8.1%) | 542 (10%) | 84 (7.0%) | 141 (7.4%) | 316 (6.9%) | 50 (7.6%) | 55 (7.3%) | 162 (11%) |
| 25-34 | 303 (21%) | 623 (23%) | 1,182 (22%) | 187 (16%) | 318 (17%) | 767 (17%) | 120 (18%) | 132 (18%) | 271 (18%) |
| 35-44 | 250 (17%) | 548 (20%) | 1,018 (19%) | 194 (16%) | 362 (19%) | 819 (18%) | 94 (14%) | 147 (20%) | 299 (20%) |
| 45-54 | 263 (18%) | 564 (21%) | 976 (18%) | 233 (19%) | 413 (22%) | 807 (18%) | 123 (19%) | 165 (22%) | 258 (17%) |
| 55-64 | 243 (17%) | 445 (16%) | 877 (17%) | 239 (20%) | 335 (18%) | 859 (19%) | 132 (20%) | 144 (19%) | 264 (18%) |
| 65-74 | 173 (12%) | 249 (9.2%) | 513 (9.7%) | 202 (17%) | 243 (13%) | 693 (15%) | 100 (15%) | 77  (10%) | 174 (12%) |
| 75-84 | 54 (3.8%) | 60 (2.2%) | 175 (3.3%) | 55 (4.6%) | 88 (4.6%) | 249 (5.5%) | 38  (5.7%) | 28 (3.7%) | 63 (4.2%) |
| 85+ | 6  (0.4%) | 9  (0.3%) | 25  (0.5%) | 11  (0.9%) | 13 (0.7%) | 58  (1.3%) | 5  (0.8%) | 3  (0.4%) | 13 (0.9%) |
| **Sex** |  |  |  |  |  |  |  |  |  |
| Female | 982 (69%) | 1,941 (71%) | 3,663 (69%) | 828 (69%) | 1,402 (73%) | 3,155 (69%) | 466 (70%) | 545 (73%) | 1,086 (72%) |
| Male | 448 (31%) | 778 (29%) | 1,645 (31%) | 377 (31%) | 511 (27%) | 1,413 (31%) | 196 (30%) | 206 (27%) | 418 (28%) |
| **Race** |  |  |  |  |  |  |  |  |  |
| AIAN | 2  (0.1%) | 7  (0.3%) | 6  (0.1%) | 3  (0.2%) | 10 (0.5%) | 8  (0.2%) | 1  (0.2%) | 1  (0.1%) | 1 (<0.1%) |
| Asian | 29 (2.0%) | 59 (2.2%) | 115 (2.2%) | 39 (3.2%) | 55 (2.9%) | 111 (2.4%) | 19 (2.9%) | 13 (1.7%) | 29 (1.9%) |
| Black | 648 (45%) | 1,135 (42%) | 2,516 (47%) | 395 (33%) | 700 (37%) | 1,624 (36%) | 306 (46%) | 373 (50%) | 848 (56%) |
| NHPI | 1 (<0.1%) | 2 (<0.1%) | 4  (<0.1%) | 0  (0%) | 2  (0.1%) | 2  (<0.1%) | 1  (0.2%) | 1  (0.1%) | 3 (0.2%) |
| White | 680 (48%) | 1,349 (50%) | 2,382 (45%) | 714 (59%) | 1,036 (54%) | 2,587 (57%) | 311 (47%) | 324 (43%) | 560 (37%) |
| Other | 70 (4.9%) | 167 (6.1%) | 285 (5.4%) | 54 (4.5%) | 110 (5.8%) | 236 (5.2%) | 24 (3.6%) | 39 (5.2%) | 63 (4.2%) |
| *^1^* n (%) | | | | | | | | | |

Table E 5: STROBE checklist of items that should be included in reports of observational studies

|  | | Item No. | | Recommendation | Page  No. | | Relevant text from manuscript |
| --- | --- | --- | --- | --- | --- | --- | --- |
| **Title and abstract** | | 1 | | (*a*) Indicate the study’s design with a commonly used term in the title or the abstract | 3 | | We conducted a retrospective EHR database study |
|  |  |  |  | (*b*) Provide in the abstract an informative and balanced summary of what was done and what was found | 3 | | Results section of the abstract |
| Introduction | | | | | | |  |
| Background/rationale | | 2 | | Explain the scientific background and rationale for the investigation being reported | 5 | | The COVID-19 pandemic prompted unprecedented changes in chronic disease self-management and healthcare systems worldwide |
| Objectives | | 3 | | State specific objectives, including any prespecified hypotheses | 6 | | We sought to characterize changes in adult asthma-related healthcare utilization during the COVID-19 pandemic in 2020 versus prior 2017-2019 and subsequent 2021-2024 |
| Methods | | | | | | |  |
| Study design | | 4 | | Present key elements of study design early in the paper | 6 | | We conducted a database study of Penn Medicine EHR |
| Setting | | 5 | | Describe the setting, locations, and relevant dates, including periods of recruitment, exposure, follow-up, and data collection | 6 | | Penn Medicine health system |
| Participants | | 6 | | (*a*) *Cohort study*—Give the eligibility criteria, and the sources and methods of selection of participants. Describe methods of follow-up  *Case-control study*—Give the eligibility criteria, and the sources and methods of case ascertainment and control selection. Give the rationale for the choice of cases and controls  *Cross-sectional study*—Give the eligibility criteria, and the sources and methods of selection of participants |  | | NA |
|  |  |  |  | (*b*) *Cohort study*—For matched studies, give matching criteria and number of exposed and unexposed  *Case-control study*—For matched studies, give matching criteria and the number of controls per case |  | |  |
| Variables | | 7 | | Clearly define all outcomes, exposures, predictors, potential confounders, and effect modifiers. Give diagnostic criteria, if applicable | 7, 8 | | Asthma Healthcare Utilization and time periods sections |
| Data sources/ measurement | | 8* | | For each variable of interest, give sources of data and details of methods of assessment (measurement). Describe comparability of assessment methods if there is more than one group | 6,7,8 | | Study design and population in the methods section |
| Bias | | 9 | | Describe any efforts to address potential sources of bias | 12 | | Limitation section |
| Study size | | 10 | | Explain how the study size was arrived at | 6 | | Supplemental Fig 1 and study population section in Methods |
| Quantitative variables | 11 | | Explain how quantitative variables were handled in the analyses. If applicable, describe which groupings were chosen and why | | 6,7 | Asthma Healthcare Utilization and time periods sections | |
| Statistical methods | 12 | | (*a*) Describe all statistical methods, including those used to control for confounding | | 8 | Statistical methods in the methods section | |
|  |  |  | (*b*) Describe any methods used to examine subgroups and interactions | | 8 | Statistical methods in the methods section | |
|  |  |  | (*c*) Explain how missing data were addressed | | 6 | Study design and population in the methods section | |
|  |  |  | (*d*) *Cohort study*—If applicable, explain how loss to follow-up was addressed  *Case-control study*—If applicable, explain how matching of cases and controls was addressed  *Cross-sectional study*—If applicable, describe analytical methods taking account of sampling strategy | |  |  | |
|  |  |  | (*e*) Describe any sensitivity analyses | | NA | NA | |
| Results | | | | | | | |
| Participants | 13* | | (a) Report numbers of individuals at each stage of study—eg numbers potentially eligible, examined for eligibility, confirmed eligible, included in the study, completing follow-up, and analysed | | 8 | Study population in Methods section | |
|  |  |  | (b) Give reasons for non-participation at each stage | | NA | NA | |
|  |  |  | (c) Consider use of a flow diagram | | 15 | Supplemental Fig 1 | |
| Descriptive data | 14* | | (a) Give characteristics of study participants (eg demographic, clinical, social) and information on exposures and potential confounders | | 8 | Study population in Methods section | |
|  |  |  | (b) Indicate number of participants with missing data for each variable of interest | | NA | NA | |
|  |  |  | (c) *Cohort study*—Summarise follow-up time (eg, average and total amount) | | 8 | NA | |
| Outcome data | 15* | | *Cohort study*—Report numbers of outcome events or summary measures over time | | 9 | NA | |
|  |  |  | *Case-control study—*Report numbers in each exposure category, or summary measures of exposure | | NA | NA | |
|  |  |  | *Cross-sectional study—*Report numbers of outcome events or summary measures | | NA | NA | |
| Main results | 16 | | (*a*) Give unadjusted estimates and, if applicable, confounder-adjusted estimates and their precision (eg, 95% confidence interval). Make clear which confounders were adjusted for and why they were included | | 9-11 | Tables 2-3: IRR with 95% CI | |
|  |  |  | (*b*) Report category boundaries when continuous variables were categorized | | 6-7 | Age groups, BMI categories, Elixhauser scores | |
|  |  |  | (*c*) If relevant, consider translating estimates of relative risk into absolute risk for a meaningful time period | | 9-11 | Percentage changes reported alongside IRR | |

Continued on next page

| Other analyses | 17 | Report other analyses done—eg analyses of subgroups and interactions, and sensitivity analyses | 11 | "Sex-, age-, and race-specific encounter trends" |
| --- | --- | --- | --- | --- |
| Discussion | | | | |
| Key results | 18 | Summarise key results with reference to study objectives | 12 | First paragraph of discussion |
| Limitations | 19 | Discuss limitations of the study, taking into account sources of potential bias or imprecision. Discuss both direction and magnitude of any potential bias | 12-13 | "Several limitations may affect the interpretation..." |
| Interpretation | 20 | Give a cautious overall interpretation of results considering objectives, limitations, multiplicity of analyses, results from similar studies, and other relevant evidence | 12-13 | Discussion section: comparison with other studies, clinical implications |
| Generalisability | 21 | Discuss the generalisability (external validity) of the study results | 12-13 | "single health system in the Philadelphia region, which may limit generalizability |
| Other information | |  | | |
| Funding | 22 | Give the source of funding and the role of the funders for the present study and, if applicable, for the original study on which the present article is based | 2 | Sources of funding: Research reported in this publication was supported by NIH..." |

*Give information separately for cases and controls in case-control studies and, if applicable, for exposed and unexposed groups in cohort and cross-sectional studies.

**Note:** An Explanation and Elaboration article discusses each checklist item and gives methodological background and published examples of transparent reporting. The STROBE checklist is best used in conjunction with this article (freely available on the Web sites of PLoS Medicine at http://www.plosmedicine.org/, Annals of Internal Medicine at http://www.annals.org/, and Epidemiology at http://www.epidem.com/). Information on the STROBE Initiative is available at www.strobe-statement.org.
